# Exploring hydration care practices in nursing homes in Brazil, a cross-sectional and focus group study

**DOI:** 10.1101/2025.11.17.25340430

**Authors:** Peter Lloyd-Sherlock, Christine Abdalla, Diane Bunn, Karla Giacomin, Philip Hodgson, Natália Horta, Nathalia Luiza Custódio, Maria Clara Santos Oliveira

## Abstract

Low-intake dehydration is prevalent among older adults, especially for those living in long-term care facilities (LTCFs). This paper presents findings from a mixed methods study of hydration knowledge and practice among LTCF managers and staff in Brazil. Drawing on an online survey with 138 respondents and focus groups in three cities, four broad analytical themes were identified: (i) gaps between theoretical understanding of hydration needs and perceived feasibility in practice; (ii) challenges in overcoming some residents’ reluctance to drink; (iii) barriers to monitoring and promoting fluid intake as part of day-to-day facility management and (iv) difficulties in sustaining interventions in contexts of resource scarcity. Our findings demonstrate a need for evidence-based interventions suited to these complex settings, to be implemented within wider frameworks of supportive public policy.

## Introduction

Regardless of the quality of water supply or sanitation infrastructure, low-intake dehydration (LID: drinking insufficiently to replace fluid losses) is prevalent among older people. A recent systematic review found one in four people aged 65 or more in non-hospital settings had LID (Parkinson et al, 2023). Causes include diminishing thirst sensation, impaired renal function and sarcopenia, which reduces the ability of the body to store water. Additional contributing factors are functional and cognitive decline, anxieties around incontinence and choking, and reduced social interaction (Hooper, et al, 2014). These risk factors are especially prevalent among older people living in long-term care facilities (LTCFs) (Yuan et al, 2022). For some older people, the loss of just two per cent of body fluid can cause physical and cognitive impairment (Edmonds et al, 2021). LID increases the risk of falls, urinary tract infections (UTIs) and other health conditions and is associated with increased risk of hospital admission and death (Hooper et al, 2021; Hamrick et al, 2020). Despite this, the condition remains under-recognized and poorly managed (Lacey et al, 2019; XXX1). There is over-reliance on clinical signs and symptoms to detect dehydration, despite strong evidence they do not work in older adults (Hooper et al, 2015; Bunn et al, 2019). The “gold standard” hydration test, serum osmolality, requires blood samples drawn by a healthcare professional and analysed in a laboratory (Volkert et al, 2022). This is not a feasible strategy for monitoring resident hydration in non-hospital settings.

Historically, numbers of LTCFs in LMICs were very small compared to high-income countries, but the sector grown rapidly over the past two decades (Wachholz et al, 2021; XXX2; Ge et al, 2025). Most facilities are privately-run, either on a for-profit or not-for-profit basis. In LMICs, official regulation and information systems are weakly developed compared to most high-income countries, with many clandestine, unregistered facilities (XXX2; XXX3). This obstructs oversight of quality assurance and daily practice.

This paper presents findings from an opportunistic mixed methods study of hydration knowledge and practice among LTCF managers and staff in Brazil. A survey was conducted in partnership with the National Front for the Strengthening of LTCFs (NF LTCF), a voluntary movement created at the start of the COVID-19 pandemic. During the pandemic NF-LTC shared information and good practice between LTCFs, developing and disseminating specific educational materials. NF-LTCF succeeded in engaging with over 7,000 LTCFs across Brazil and has been credited with contributing to relatively low levels of COVID-19 mortality in facilities (XXX4).

As the pandemic abated, NF-LTCF looked to share knowledge and improve practice in areas less specifically related to COVID-19. NF-LTCF had links with UK researchers who had piloted interventions to improve hydration practice in local LTCFs. Since there were no similar examples in Brazil, hydration was identified by NF-LTCF as a priority concern. Initial activities included exchanges between Brazilian and UK experts and the modification of an intervention trialled in UK LTCFs to suit the Brazilian context (XXX5). Additionally, NF-LTCF developed an online survey of members to assess their knowledge, practice and degree of concern about resident hydration. This was followed by focus group discussions (FGDs) with LTCF managers and staff in the cities of Belo Horizonte, Rio de Janeiro and Salvador de Bahia. This paper reports the findings from the online survey and focus groups and identifies key issues for policy and practice.

## Materials and methods

The study was subject to ethical review by the University of XXX (XXX) and the XXX University (XXX). It applied a sequential mixed methods design, including an online questionnaire and focus groups to explore current understandings of hydration amongst senior LTCF staff (Creswell et al, 2003). Findings from each study were analysed separately before combining for final interpretation.

In all countries, there are challenges to researching care practice in LTCFs, including recruitment, participation and reliable reporting (XXX1). In many facilities, senior staff have limited availability to participate in research and express understandable concerns about researchers disturbing care routines or unsettling more vulnerable residents (Lam et al, 2018). These barriers were partly mitigated in our study, as the research team included members of NF-LTC who enjoyed a high degree of trust and goodwill following the COVID-19 pandemic. Nevertheless, research did not take place in the facilities themselves and relied on reported behaviour rather than direct observation.

The research team developed a short, structured questionnaire for LTCF managers or other senior staff, remotely accessible via Google forms (Appendix 1). The form included closed questions on knowledge about hydration, challenges in maintaining resident hydration and any strategies they applied. NF-LTCF presented the proposed study to its members in a live webinar in December 2023 and a link to the form put on its website (XXX).

Survey questions were based on those developed and used by the UK research team and adapted for the Brazil context. Participants were reassured that participation was voluntary and would neither be rewarded nor reimbursed. The questionnaire included an invitation to join follow-up focus group discussions (FGDs) to explore the same issues in greater depth (topic guide available in Appendix 2). Data collected through the online survey were analysed descriptively, as the sample was not sufficiently large for multivariate analysis.

The offer to join FGDs was accepted by 121 respondents, of whom 21 were located in Rio de Janeiro, Belo Horizonte or Salvador. They were invited to a FGD in each city by email and a follow-up phone call and eleven went on to participate in person. Where provided, the main reason for non-participation was the unavailability of a senior staff member to attend at the specified time and place. The 11 participants included five managers, three nurses, two nutritionists and one nursing assistant (Table 1).

**Table 1.** Focus group discussion participants.

| Informant code | City | Type of LTCF | Role |
| --- | --- | --- | --- |
| RJ1 (FP) | Rio de Janeiro | Private | Nurse |
| RJ 2(FP) | Rio de Janeiro | Private | Nursing assistant |
| RJ 3(FP) | Rio de Janeiro | Private | Manager |
| RJ 4(FP) | Rio de Janeiro | Private | Nurse |
| RJ 5(FP) | Rio de Janeiro | Private | Nutritionist |
| RJ 6 (FP) | Rio de Janeiro | Private | Nurse |
| SV1 (FP) | Salvador | Private | Nurse |
| SV 2(NFP) | Salvador | Not-for-profit | Manager |
| SV 3(NFP) | Salvador | Not-for-profit | Manger |
| BH 1 (NFP) | Belo Horizonte | Not-for-profit | Manager |
| BH 2 (NFP) | Belo Horizonte | Not-for-profit | Nutritionist |

The FGDs took place in March 2024 and lasted around two hours. Participants were provided with information about the study and completed and signed informed consent forms before joining the discussions, which were recorded. Other than refreshments, participants were not provided with payment or direct incentives, although there was an opportunity to share knowledge and experience about the research issue. X1, who speaks fluent Portuguese, facilitated the FGDs, and all groups were co-facilitated by two or more members of the Brazilian authorship team, either in-person or remotely. The FGDs encouraged interactive dialogue between participants, including areas of dissonance or disagreement.

Qualitative data analysis followed the MIRACLE narrative framework for thick description, applying an authentic, person-centred and relational lens, situated within specific institutional and cultural settings (Younas et al, 2023). Recordings were transcribed verbatim by X2 and X3, and analysed thematically by X4, X1, X2 and X5. FGD content was coded, categorised and key sub-themes were identified independently by Brazilian members of the research team and by X1 prior to team discussions. Analytic sub-themes were then expanded and organised into broader themes. Differences in identified themes and interpretation of findings were then reconciled. Direct quotations from FGD participants are presented in the text, selected on the basis of authenticity and argument criteria (Lingard and Watling, 2021). Quotations were translated by X1, a native English speaker, and agreed with other members of the authorship team. Original language versions are available on request.

Although mixed methods add interpretative value to studies, they create complexity for the coherent analysis and presentation of research findings (O’Cathain et al., 2010). The FGDs were informed by preliminary analysis of the survey data. However, some themes emerging in the FGD narratives, such as monitoring fluid intake, had not been anticipated by the online survey. Consequently, research findings are organised by broad qualitative themes and linked to corresponding survey data where this is available. The presentation of findings follows COREQ guidelines for qualitative research which are presented in Supplementary File 1 (Tong et al, 2007).

## Results

At the time of paper submission, 145 LTCF staff had completed the online survey. Of these, seven had completed it incorrectly and were excluded from the study. Table 2 presents general information about 138 facilities, along with responses to questions related to this study. The geographical distribution of facilities reflects that reported by other studies (Guimarães et al, 2024), with nearly half located in the South-East. Other LTCF characteristics, such as profit or non-profit status and size, were also broadly representative of national trends (Wachholz et al, 2021). Non-profit homes included those which were directly managed by state agencies and those run by philanthropic organisations.

**Table 2.** Profile of LTCFs participating in the online survey and selected responses (n=138).

|  |  | Total |
| --- | --- | --- |
| Region | North | 1 |
|  | North-East | 21 |
|  | Centre | 15 |
|  | South-East | 82 |
|  | South | 19 |
| Funding | For-profit | 46 |
|  | Not for-profit | 92 |
| Number of residents | 1-15 | 19 |
|  | 16-30 | 56 |
|  | 31-50 | 41 |
|  | 51+ | 22 |
| Knowledge | 1 litre or less per day | 7 |
|  | 1.1 to 2 litres per day | 100 |
|  | Over 2 litres a day | 30 |
|  | Don't know | 1 |
|  | Maintain organism physiology | 96 |
|  | Disease prevention | 81 |
|  | Other | 5 |
| Challenges | Refusal or reluctance to drink | 116 |
|  | Specific health conditions | 35 |
|  | Demands on staff time | 13 |
|  | Lack of awareness of the importance of hydration | 59 |
|  | Other | 14 |
| Strategies | Offer more to drink | 63 |
|  | Use drinking bottles and monitor intake | 22 |
|  | Protocols and structured care routines | 36 |
|  | Offer alternative drinks and vary how they are provided. | 45 |
|  | “Special drinks” offered to residents with specific needs | 73 |
|  | Training and awareness raising | 27 |
|  | Specific strategies to improve staff hydration | 74 |
|  | Additional strategies in heat wave. | 97 |

**Table 3.** Themes and sub-themes with Illustrative quotations.

| Main Theme | Sub-theme | Example |
| --- | --- | --- |
| **Knowledge** | Quantity and viability | At least two litres. But it's an enormous challenge - almost mission impossible. SV2 (NFP) |
|  |  | I'd say we estimate 35 millilitres per kilo, but of course normally they won't even drink half of that. BH1 (NFP) |
|  | Symptoms and warning signs | We keep an eye open for dry skin or dry mucus. Or if their urine has a strong smell or there isn't much of it, when we change their pads. RJ3 (FP) |
|  |  | Usually, they start to get mentally confused. For residents with dementia, we know what they are normally like, so if their behaviour changes, something not like them, we start to wonder if they are dehydrated. BH2 (NFP) |
| **Resident reluctance** | Need for persistence. | You have to keep offering them drinks every day and be persistent with it. Doing that, you should get good results. RJ5 (FP) |
|  | Some residents are hard work. | I see staff insist more with some residents than with others. They have residents they feel closer to than to others. Some are really hard work. They call for the caregivers all day long and it wears them out. BH1 (NFP) |
|  | Coercion. | Where I work, I always say you don't <u>offer</u> them water or ask if they want some. You give them the cup and they have to drink it. You can't ask them first. It's the same thing with food. If you ask them if they want a drink, they'll just tell you they don't, that they'd already just had something. RJ6 (FP) |
|  | Reduce toilet need. | There's one resident who is on blood pressure pills and so she has to keep going to the toilet all day. She thinks if she drinks a lot of water she will have to go even more. And so, she hardly drinks any water at all. And she should really be drinking twice as much because her medicine makes her wee so much. RJ4 (FP) |
| **Daily management** | Bottles. | They all have little bottles in their rooms, and I tell them "You need to drink so many little bottles a day." BH1 (NFP) |
|  |  | I get WhatsApps at the end of each shift to report on how things have gone, with photos of the bottles and the amount of water residents have drunk during the day. (RJ2 FP) |
|  | Monitoring charts and books | I put up charts for daily hydration saying how much each person should drink a day. And I keep reminding the staff through the day that they need to get to at least one and a half litres. If they don't manage that, the night shift has to make up the difference. RJ3 (FP) |
|  |  | Our staff have a booklets, kept in residents' rooms. They have to update them and show them to family members too. They put in things like when the person ate or had a snack... Things like: 8 o'clock breakfast, 9.30 this type of medication. We only do this for the residents who are quite care dependent. But the booklets don't say anything about amounts. Just that the person drank some water, but not how much. BA2 (NFP) |
|  |  | You need to keep reminding staff. I think it's good to explain to them about all the problems dehydration can cause. I really scare them about what dehydration can do to people, and they get very concerned. "Oh my god! I have to keep giving them water!" Staff get really vigilant about this, and we keep an eye on the drinking bottles and on how much has been drunk after meals. RJ1 (FP) |
| **Special strategies** | Different drinks | When we make flavoured water they drink it, even if they're not thirsty. Because they see that it's different. It looks more interesting. BH2 (NFP) |
|  |  | In our place flavoured water doesn't work that well, so we try to offer them juice. And there's always tea available. We put teabags into jugs. We make a lot of jelly too, either set or in liquid form. BH1 (NFP) |
|  |  | Today is Women's Day, so we made two jugs of passion fruit juice. And so, as well as their usual breakfast, they all had half a cup of juice. It's something different for them.<br>BH2 (NFP) |
|  | Unhealthy drinks | We know you shouldn't really swap flavoured water for regular water. But it's something we learned to do so that residents take on more fluid. RJ 5 (FP)<br>We offer them any kind of drink they want. We only refuse to serve sodas. BA2 (FP) |
|  | Cost | It's very expensive... a litre of coconut water costs 20 *reais* in the greengrocers.... When we had that heatwave, I asked family members to give us coconut water. I knew it was important to have it... But then there are families and there are families, if you know what I mean. RJ3 (FP) |
|  | Drinking as social activity | When residents are together, and someone starts to drink water the others will say "I want some too". BA1 (FP)<br>When we do exercises this sometimes makes them want to drink more water. BA2 (FP) |
|  | Reminders | I put posters up in strategic spots about how important drinking water is for everyone. BH1 (NFP) |

Four broad analytical themes were identified from the survey and FGD data: (i) knowledge of hydration needs; (ii) residents’ reluctance to drink; (iii) daily management of hydration and fluid intake (“business as usual”) and (iv) special strategies to promote drinking (“Going the extra mile”).

### Knowledge of hydration needs

Predictably, all responses to the questionnaire stated residents’ hydration was an important issue which affected their physical health status (Table 2). Most respondents viewed hydration as necessary for disease prevention, with specific reference to conditions such as UTIs. Similar comments were made in the FGDs:

> *We noticed a very strong odour of urine and some traces in the urine that look like faeces… It’s an older lady who was hospitalized 3 times due to UTIs, 3 months on the trot. She’s one of our residents who drinks the most water, although it’s still very little. So, imagine the others! This woman already had kidney failure due to excessive use of antibiotics.* BH1 (nfp)

In this case and in others, FGD participants referred to a number of indicators or warning signs often thought to be associated with dehydration. No mention was made of more reliable or objective assessment from blood samples.

The recommended daily fluid intake requirement for adults of all ages is 1.6 litres for women and 2.0 litres for men (Volkert et al, 2022; Gonçalves et al, 2019). The survey asked about the recommended daily volume of fluid intake for older people, without differentiating by sex and 107 respondents recommended 2 litres or less (Table 2). In the FGDs the same question was framed more openly, without specific response categories. Participants in all three groups provided answers exclusively in terms of litres (though no metric was mentioned in the question), and some specified amounts in relation to body weight or the occurrence of heat waves.

> *2 to 3 litres a day, on a normal day… I guess when it’s a hot day, maybe double that… Residents lose more liquid on hot days and if you don’t double the amount, they are going to get dehydrated.* RJ2 (fp)

Participants were then asked to relate these litre values to the volume of fluid contained in the drinking vessels (cups, flasks etc.) residents most frequently used. Several found this difficult, suggesting a disconnect between their theoretical knowledge and daily practice. More generally, FGD participants reported that fluid intake guidelines often represented an unfeasible target.

> *On a normal day, we give them 8 cups. Residents have a lot of difficulty drinking water, and we need to be open about these problems. In an ideal world, you would say 8 cups… And, of course, not all of them drink the whole cup.* RJ1 (fp)

### Residents’ reluctance to drink

The survey asked about the most important challenges in keeping residents hydrated (Table 2). The most frequent response (116) was the refusal of some residents to drink sufficiently. FGD participants agreed that resident hydration sometimes required considerable time, patience and persuasive skill on the part of staff, and some referred to coercive practice.

> *We need to get to know what each resident is like. Some of them like to be praised by saying “Go on, drink some water so your skin looks prettier. Your daughter’s visiting today and your skin looks so dry.” And we are careful about how we select care staff. They need to be really patient, attentive and charismatic, because some care staff show up with a sulky face and the residents don’t want to know them.* SV1 (fp)

> *We had a woman who didn’t want to open her mouth to drink water. And so we tried everything we could … [respondent pinches her nose and opens her mouth].* RJ1 (fp)

> *We sometimes offer alcohol-free beer and the residents with dementia don’t realise it’s zero alcohol. We put it in a different type of cup, inside a plastic beer cooler, and then they drink it as if it’s the same beer they have been drinking all their lives* SV1 (fp)

FGD participants agreed many residents sought to reduce fluid intake to limit the frequency of toilet visits or to manage incontinence. Some care staff saw this as residents being difficult, rather than due to genuine anxiety or a physiological issue.

> *Some residents are too lazy to get up to go to the bathroom after drinking. “Oh, I really don’t want any water because I have to go to the toilet all the time.” You give them a cup and they throw half of it away. Too lazy to walk!* SV2 (nfp)

> *Then there are those care workers who don’t want to provide water to residents because they don’t want the bother of taking them to the toilet. These are issues for the care home management to deal with.* SV1 (fp)

In another instance, there was recognition of genuine fear associated with incontinence, beyond the normal anxieties associated with loss of dignity and stigma:

> *We have a woman who told us that her mother used to hit her when she wet the bed. And so, she doesn’t want to drink water. She is so afraid of wetting her bed again.* BH1 (nfp)

FGD participants agreed that managing incontinence by limiting drinking was undesirable. However, some acknowledged that reducing the frequency of toilet visits or pad changes could ease staff workload and cost pressures. The cost of incontinence pads was a major consideration, both when it was met by the facility or when they were paid for by residents’ families.

> *Residents call a carer to help them go to the toilet, but sometimes they hold on for a long time. If they need a pad change before going, the “girls” will bring them it. Just to give you an idea: we get through about 1,300 adult pads a week.* BH2 (nfp)

> *We took in a resident who had a home carer before and only used 2 pads a day. And when she got here, we were using 6 to 8 a day, and so the family complained that they were spending too much on pads. But it was just that we were hydrating them at the right times of day… And sometimes the residents themselves tell us: “Selma [name of manager], my son is complaining about the cost”. But if we don’t change the pads, the patient will get an infection. “Alright then; you talk to my son”. And we talk to him….* SV1 (fp)

### Daily management of hydration and fluid intake (“business as usual”)

A minority of survey respondents (36) referred to applying protocols about the provision of drinks at certain times of day and/or recording intake (Table 2). Some FGD participants commented that residents hydrated by tube were the most easily monitored and managed.

> *We try to give them water during lunch every day, and there are fixed times in the morning, like between their showers and elevenses, as well as between elevenses and lunch, and in the afternoon… The residents hydrated by tube always get fluid at the right times and the amount indicated by the nutritionist: there’s no problem with them. But there are residents who you have to really insist with to drink water and to eat, who need more time to eat. They give us more work. They’re usually the ones with dementia.* BH2 (nfp)

The survey did not ask how resident fluid intake was monitored, but this emerged as a key concern in the FGDs. Several participants had attempted to develop systems for reporting and monitoring intake, although this was considered less necessary for residents who could get their own drinks.

> *I think we just have to trust that our staff are providing drinks. We have registers that staff sometimes fill out in an automatic way or sometimes don’t fill out at all… There are residents who can get their own water from the drinking station without any help and there’s no way to keep track of how much they drink. Some residents are always drinking, more than is necessary, they just keep on drinking.* BH2 (nfp)

> *Our staff have a booklet that they have to update and even share with the families. They put down the time of meals and things like that… At 8 o’clock they had breakfast, at 9.30 they took this pill… We just do this for the more dependent residents. But the booklet doesn’t have anything about amounts. It says whether they drank some water, but not how much they drank.* SV2 (nfp)

Tap water in Brazil is not potable and so LTCFs usually provide filtered water at drinking stations. Only 13 survey responses referred to pressures on staff time as a key challenge for ensuring resident hydration. However, in the FGDs, this issue was given more emphasis, particularly in not-for-profit LTCFs.

> *For 46 residents I should always have eight care staff present. But sometimes maybe three carers are taking residents for health visits, and some are busy bathing residents. And so, I’m just left with two or three staff for day-to-day stuff. In the kitchen I have just one cook and a helper to prepare meals and drinks for everyone.* BH2 (nfp)

FGD participants commented that resident hydration should be the responsibility of all staff, not just caregivers, and that communication was the key to enabling a team approach. However, they disagreed about how motivated staff were to persist with drinking, especially in facilities with lower staff to resident ratios.

> *I think it’s all about communication. There was an issue with some of our kitchen staff: some of the cups were being returned to the kitchen with juice still in them. And it was only when management became aware that we were able to discuss it with staff. If they’d told us about it sooner, we could have dealt with it faster.* BH1 (nfp)

Participants also discussed the frustrations and challenges of trying to help reluctant residents to drink:

> *It’s quite comical sometimes. A member of staff will come up to me saying, “-[name], I’ve tried everything and I’m at my wit’s end. I tried putting sweetener in their water, I tried juice with a straw, but they just won’t drink.” They bring me this feedback; they don’t simply ignore the problem.* RJ4 (fp)

### Special strategies to promote drinking (“Going the extra mile”)

In the survey, 63 respondents mentioned increasing the amount of fluid offered as a key strategy and 45 referred to increasing the variety of drinks how they are offered (Table 2). All FGD participants referred to these strategies and considered them useful. Several participants mentioned that social activities could encourage drinking, but that these were difficult to organise in contexts of staff shortages and rigid routines.

> *We had this project where the residents would make their own ice lollies… But then we never had the time. We are short of staff after someone had to leave, and so things are always crazy and we never have the time for it.* BH1 (nfp)

“Special drinks for older people” (usually understood to be thickened drinks or nutritional supplements) were offered by 74 LTCFs. In less well-resourced facilities, the choice of drinks was usually limited to water or sugar-flavoured water. In Brazilian care homes, water is the most commonly provided drink, rather than hot beverages such as tea. The survey reported 10 LTCFs, including 6 out of 7 state-funded ones, provided residents with unfiltered (and therefore non-potable) tap water, whereas in facilities with more resources a wider range of drinks was available. One FGD participant raised health concerns about flavoured water, and there were different views about the suitability of sodas and alcoholic drinks due to the sugar content and impact on oral health, and perhaps other conditions such as diabetes.

> *We had a visit from a public health regulator looking inside all the fridges… She asked “Do you allow your residents to drink soda?”. And I said “No, we don’t.” But then a resident heard us and said her nephew had brought her some soda… She’s spent her whole life drinking soda, so how can we just take it off her?* SV3 (nfp)

> *We’ve tried lots of things, even alcohol-free beer. Many of our residents were admitted here because they had alcohol problems, so we are strict about our no-alcohol policy.* BH2 (nfp)

Over half of LTCFs surveyed said they adopted additional strategies to promote resident hydration during heat waves (Table 2). The most common ones discussed in the FGDs were offering residents ice lollies and drinks that were not usually provided, such as coconut water.

> *We only offer coconut water when we get it from Mesa Brasil [a national food bank]. Other than that, coconut water is just too expensive for us.* BH2 (nfp)

Lack of staff awareness was identified as a key challenge by 59 survey respondents and 27 referred to training and awareness raising as key potential strategies (Table 2). However, none of the FGD participants were aware of any specific resources related to hydration training.

> *Nobody here has ever thought about doing that [specific staff training or support for resident hydration]… We’ve never done any kind of course, any kind of talk about that sort of thing.* BH2 (nfp)

Specific strategies to promote hydration of staff themselves were applied by 74 of the survey respondents (Table 2). These included promoting staff awareness that hydration was not just important for residents and ensuring that water and other drinks were easily accessed during work shifts.

## Discussion

This mixed-methods study explored attitudes and practices relating to resident hydration in Brazilian LTCFs. Despite the relatively large number of survey respondents, the data cannot be taken as potentially representative of all LTCFs in Brazil, which currently number over 8,000. The voluntary, self-selecting nature of the survey is likely to have generated a degree of response bias. As such, the survey finding that all respondents considered resident hydration to be an important issue, may not reflect attitudes in other facilities. Likewise, LTCF staff participating in the focus groups were already sufficiently engaged with the issue that they felt it worth their while to attend. Additionally, members of the study team did not request access to LTCFs or to speak with residents. There is a clear need for further research to address this study’s limitations.

Notwithstanding these caveats and other limitations of the study design, this is the first study to explore attitudes and knowledge about hydration care in LTCFs either for Brazil or Latin America more generally. Several useful insights were generated. Despite their general interest in the issue of hydration, a significant proportion of respondents under-estimated older people’s daily fluid intake needs. It follows that there is a need to educate senior LTCF staff about these requirements. This should recognise that international guidelines do not refer to contexts of very high temperatures, when fluid intake needs will be considerably greater (Brennan et al, 2020). This is increasingly important in Brazil, which experienced a trend of increasingly frequent and severe extreme heat events in recent decades (Bitencourt et al, 2019). As such, there is a need to develop separate hydration guidance about extreme weather events and ensure it is communicated to LTCFs.

The survey and FGDs indicate that educating staff about resident hydration needs is unlikely to bring about practice change, unless it is accompanied by interventions addressing wider and more complex sets of barriers. Many of these relate to workforce and resource constraints. The FGDs reveal the considerable time, patience, ingenuity and skill required to assist some residents to drink. As in high-income countries, this task occurs in contexts of low staffing ratios, rigid daily routines and financial constraints, and it has to be balanced against other demands on staff time (Gasper, 2011; Cook et al, 2019b).

FGD participants noted challenges related to incontinence and use of toilets, and that staff sometimes resorted to unethical strategies to get residents to drink. Both examples of reported practice judged by the authorship team as potentially unethical are presented. These include a case of deception. The ethicality of this practice when addressing a fundamental need (such as hydration) for an older person with cognitive impairment remains highly contested (Dresser, 2021; Murray et al, 2024). The other case consists of a caregiver pinching a resident’s nose, presumably so they would open their mouth to accept fluid. There is no ethical justification for this practice, although the informant’s openness indicated they were unaware of this. The research team informed the informant that this was unethical. No further action was taken for several reasons. First, the caregiver’s gesture was not captured on the FGD audio recording. Second, other studies have noted that official responses to complaints, such as removal of a resident to a different facility, may create more harm than allowing them to remain, once concerns have been raised (Donovan and Regehr, 2010).

The FGDs also revealed challenges in monitoring levels of resident fluid intake. Similar findings, including a tendency to record what is offered rather than what is consumed, have been reported for other countries, as well as imperfect application of fluid intake charts (Cook et al, 2024).

In the light of these difficulties, it is useful to consider what is feasible (or seen as feasible) by LTCF staff and managers. The FGDs reveal a large gap between the “ideal world” recommended number of litres and what participants considered possible in daily practice. Drawing attention to gaps between recommendations and practice may simply generate a sense of failure and of unrealistic demands on staff, and may promote unethical practice, with staff coercing reluctant residents to drink. It may be more effective to strike a balance between ideal goals and more achievable narratives of practice change. This corresponds with wider evidence about the effectiveness of more incremental quality improvement strategies in LTCFs (Pimentel et al, 2020; Peryer et al, 2022).

Consideration should be given to the role of regulatory agencies in promoting and ensuring LTCF resident hydration. To date, hydration remains neglected by public agencies with responsibility for LTCFs in Brazil. For example, the Ministry of Health has published detailed quality standards for LTCFs, including the need to offer older people culturally acceptable food six times a day, but makes no reference to drinking or hydration (Ministry of Health, 2025). Including hydration in these standards is essential, but it should be done in a way that facilitates improvement rather than simply drawing attention to shortfalls. Current approaches to LTCF regulation in Brazil place a strong focus on policing ideal standards, rather than developing partnerships with providers to improve practice (Ministry of Women, Family and Human Rights, 2021). Significant numbers of LTCFs in Brazil are unregistered in order to avoid regulatory oversight (XXX3). A further concern for LTCFs may be litigation from family members if unrealistic standards are not met.

Relatedly, regulatory agencies apply a strong public health lens, treating LTCFs as healthcare institutions more than as older people’s homes. This is reflected in policies to ban sodas and low-alcohol beverages on health grounds. There is strong evidence about the harmful effects of these products and that they are particularly severe for older people (Sun et al, 2023). However, it is essential to consider wider issues about older people’s personal freedoms, as well as the balance between potential harmful effects and benefits due to increased hydration.

Policies and interventions to promote hydration should recognise that sustained practice change must come from within LTCFs rather than be imposed from above (Windle et al, 2023). The survey and FGDs provided examples of strategies, such as increasing the variety of drinks being provided within limits of affordability. FGD participants referred to creative approaches which resonate with evidence from high-income countries about the value of promoting drinking as a social and enjoyable activity for residents as well as staff (Cook et al, 2019a; Cook et al, 2024; XXX6). There is evidence from other countries that dehydration is prevalent among health professionals (El-Sharkawy et al, 2015). As such, hydration strategies may be more acceptable if they take a whole of institution approach, acknowledging shared needs and collective responsibilities. Related interventions in high-income countries may provide useful insights for Brazil, but they should be mindful of substantial contextual differences and the need for interventions to be developed from within facilities (NICE, 2018; Bruno et al, 2021). Similar hydration interventions have not been systematically tested in LTCFs in Brazil or other LMICs, although a small pilot was developed and trialled in parallel with this study (XXX5).

Regardless of their design and implementation, interventions will have limited sustained impact unless accompanied by additional resourcing and other forms of support. Service improvement in this area, as well as with reference to issues such as nutrition, hygiene and infection management, is often challenging in LTCFs, and it requires multi-dimensional strategies to address complex causes (Lavallée et al, 2018). Low staffing ratios, and difficulty affording pads or more favoured drink options demonstrate the need for higher levels of funding for LTCFs both by the state and by residents’ families. There is a powerful case for greater state support, in terms of both the human rights of residents and economic arguments (such as reduced hospital admissions for conditions caused or exacerbated by dehydration). Without greater recognition and support for LTCFs, there will be little progress in addressing resident dehydration or other practice challenges.

## Data Availability

The data that support the findings of this study are available on request from the corresponding author. The data are not publicly available due to privacy or ethical restrictions.

## Acknowledgements

The authors gratefully acknowledge the support provided by the Institutional Scientific Initiation Scholarship Program (PIBIC) of Brazil’s National Council of Scientific and Technological Development (CNPq).

## Supplementary file. Consolidated criteria for reporting qualitative studies (COREQ): 32-item checklist

Developed from:

Tong A, Sainsbury P, Craig J. Consolidated criteria for reporting qualitative research (COREQ): a 32-item checklist for interviews and focus groups. *International Journal for Quality in Health Care*. 2007. Volume 19, Number 6: pp. 349 – 357

## STUDY TITLE: Avoiding dehydration among older people in Brazilian long-term care facilities

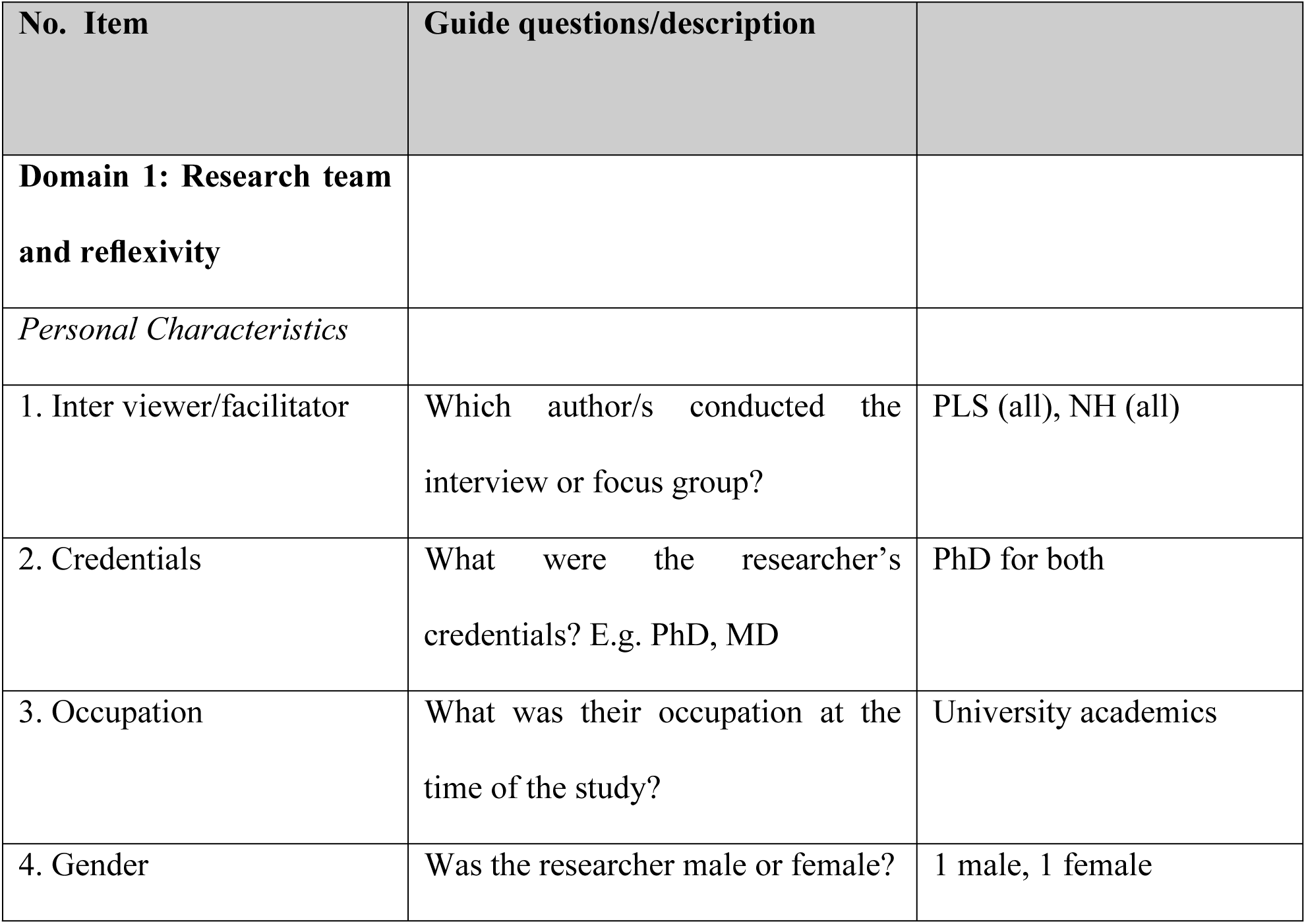
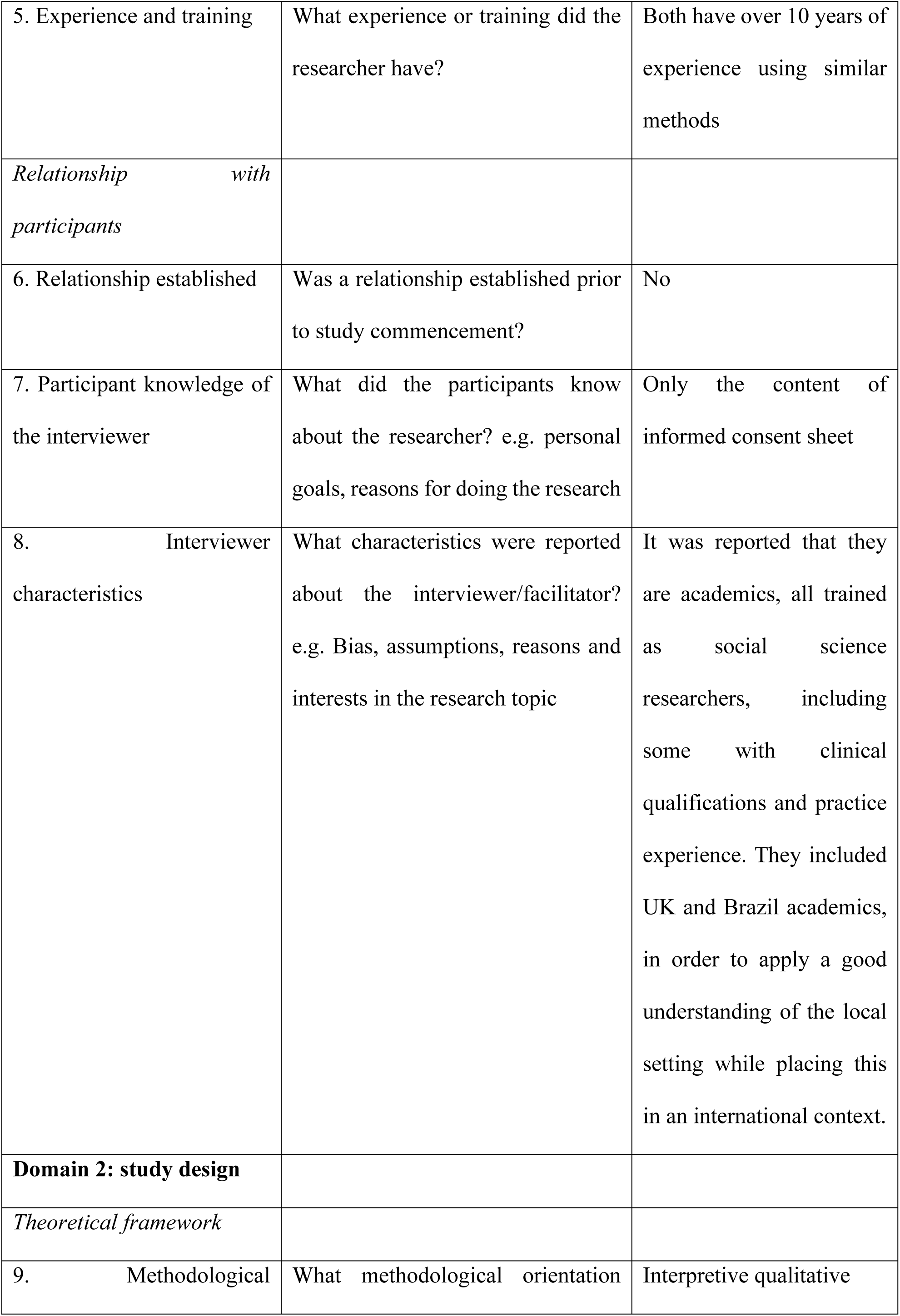
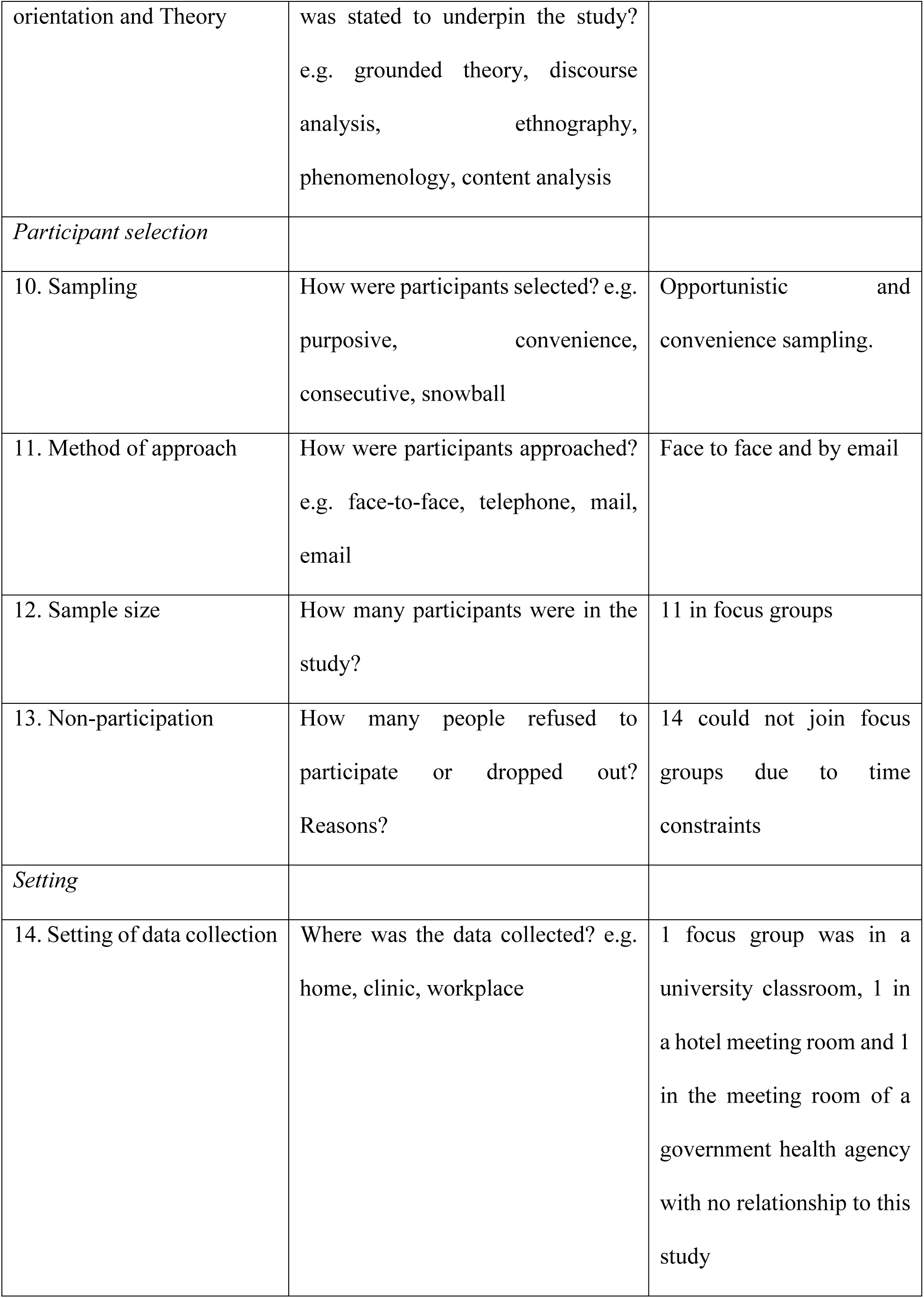
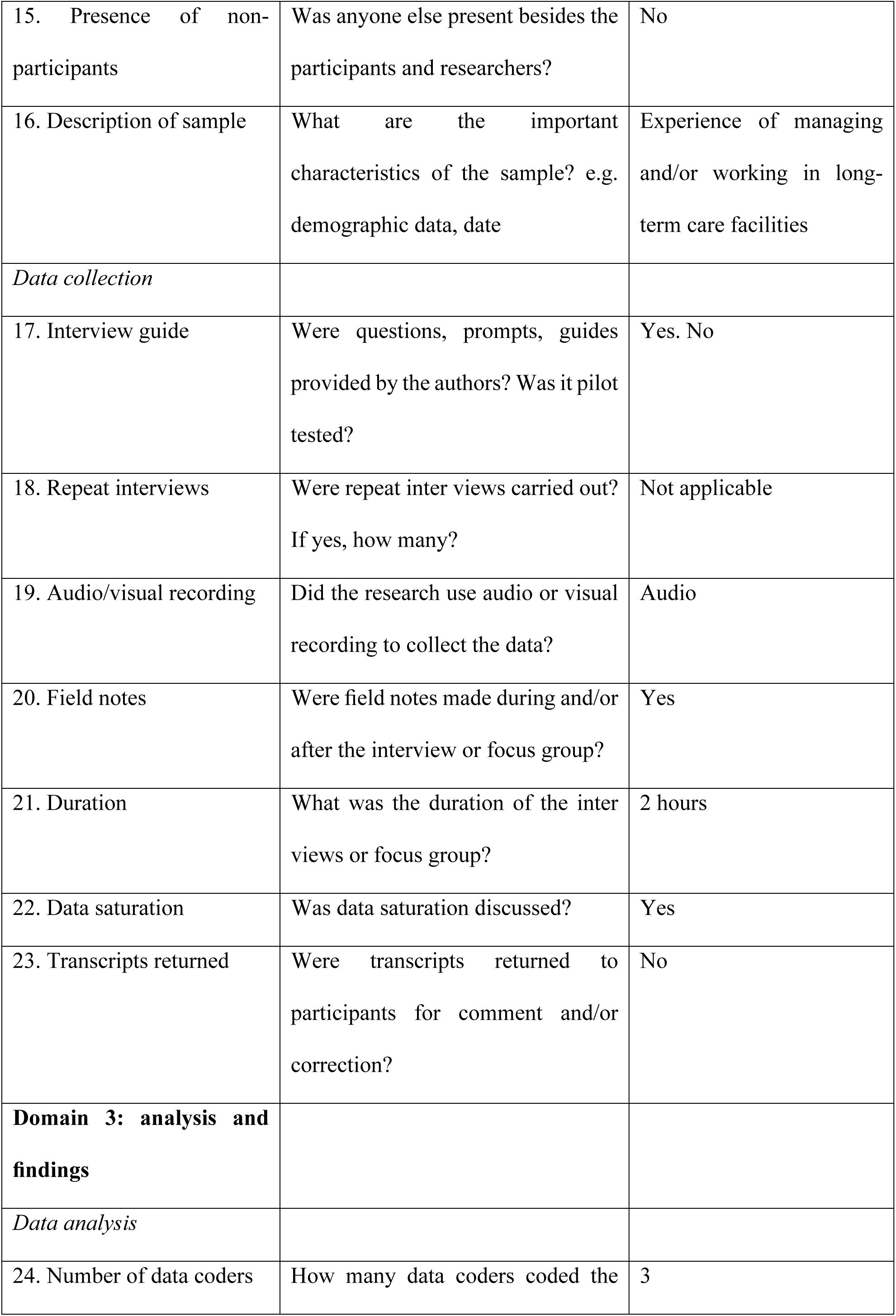
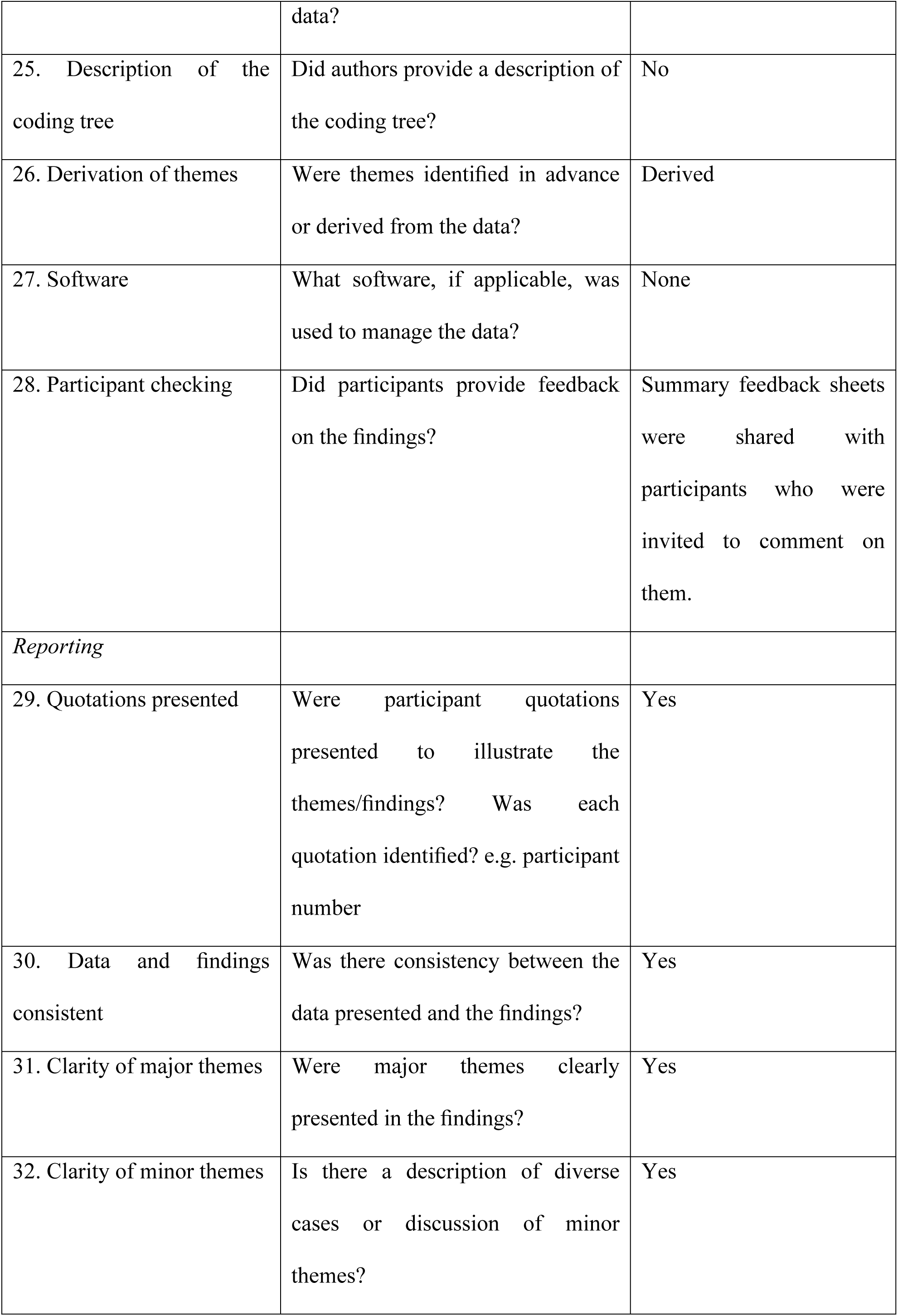

## Appendices

## Appendix 1 Online survey form

1. Name of the LTCF:

2. Nature: ( ) Public ( ) Private for-profit ( ) Private non-profit

3. Total number of residents:

4. Number of residents by degree of dependency: Grade 1; Grade 2; Grade 3

5. Do you have a nurse in your team of professionals at the LTCF? ( ) Yes ( ) No

6. Do you have nursing assistants and/or technicians on your team? ( ) Yes ( ) No

7. Do you have caregivers for older people on your team? ( ) Yes ( ) No

8. In general, how much do you think residents (men and women) need to drink for day?

( ) 0.5 liters ( ) 1 liter

( ) 1.5 liters ( ) 2 liters

( ) more than 2 liters

( ) I don’t know how to answer

9. What kind of drinks do you offer at LTCF? You can check more than one option:

Tap water Yes/No

Filtered water Yes/No

Coffee Yes/No

Tea Yes/No

Juice Yes/No

Milk Yes/No

Porridge Yes/No

Other Yes/No

Special drink for seniors Yes/No

10. Do you have a water filter/drinking fountain in your LTCF? ( ) Yes ( ) No

11. If so, is it in an accessible place for residents? ( ) Yes ( ) No

12. Please point out two (2) reasons why you think hydration is important for older people:

13. Please point out 3 (three) problems/challenges you face in order to maintain the hydration of residents in the LTCF routine:

14. What actions does your LTCF take to ensure the hydration of residents?

15. Do you have any action to improve the hydration of LTCF professionals? ( ) Yes ( No)

16. Would you be interested in training materials to help improve your hydration of residents? ( ) Yes ( ) No

17. What needs do you experience to improve the hydration of residents?

18. Would you be interested in participating in a group discussion on this topic? ( ) Yes ( ) No

## Appendix 2 Topic guide for focus group with LTCF managers/professionals

General information about the LTCF.

Public, private, philanthropic
Number of residents: total and by degree of dependency
Participant information.

Training, time working in the LTCF
How important for you is hydration of your residents?

Reasons why you think hydration is important for older people.
About the residents’ hydration routines:

How much do you think residents need to drink per day?
What do you usually offer residents to drink and how?
Do you offer them a choice of drink, temperature of it, etc.?
What kind of drinking vessel do you use?
Do some residents have more problems than others?

If so, what are they?
What are the main problems to ensure the hydration of residents?

And how do you try to solve them?
How do you help residents better hydrate and cope with related difficulties?
What specific actions does your LTCF take to improve the hydration of residents?
Do you have any strategies to improve the hydration of your staff?
Would you be interested in training materials to help you improve hydration of your residents?

## Notes

### Competing Interest Statement

The authors have declared no competing interest.

### Author Declarations

The Ethics Review Committees at the University of East Anglia, UK (approval: ETH2324-0846), and the Pontifícia Universidade Católica de Minas Gerais, Brazil (approval: CEP PUC Minas 6.421.224), approved our interviews on October 10, 2023.

